# Quit attempts and tobacco abstinence in primary care patients: long-term follow-up of a pragmatic, two-arm cluster randomised controlled trial on brief stop-smoking advice (ABC versus 5As method)

**DOI:** 10.1101/2021.03.15.21253613

**Authors:** Sabrina Kastaun, Wolfgang Viechtbauer, Verena Leve, Jaqueline Hildebrandt, Christian Funke, Stephanie Klosterhalfen, Diana Lubisch, Olaf Reddemann, Tobias Raupach, Stefan Wilm, Daniel Kotz

## Abstract

We developed a 3.5-h-training for general practitioners (GPs) in delivering brief stop-smoking advice according to different methods (ABC, 5As). In a pragmatic, cluster randomised controlled trial our training proved effective in increasing GP-delivered rates of such advice. In this follow-up analysis we examined the effect of ABC and 5As on patient-reported quit attempts and point prevalence abstinence at weeks 4, 12, and 26 following GP consultation.

Follow-up data were collected by questionnaires delivered to 1,937 smoking patients recruited before or after the training in which 69 GPs participated. At week 26, ∼70% of the patients were lost to follow-up. All 1,937 smoking patients were included in an intention-to-treat analysis. Multiple imputation was used to impute missing outcome data of each follow-up and few missing data of potential confounders.

While the receipt of brief GP advice compared to no advice was associated with a two-fold increase in patients’ attempts to quit at each follow-up and abstinence at week 26, quit attempts and abstinence rates did not differ significantly from pre- to post-training or between patients from the ABC versus the 5As group.

We previously reported that our training increases patient-reported rates of GP-delivered stop-smoking advice. The present follow-up analysis did not show that patients more often attempted or achieved to quit when they received advice from a trained vs. untrained GP. Future training studies should take contextual factors into account – such as access to free evidence-based cessation treatment – which might hamper the transfer of GPs’ effective advice into patients’ behaviour change.

**TRIAL REGISTRATION:** German Clinical Trials Register, prior to the first patient in (DRKS00012786, www.drks.de/drks_web/setLocale_EN.do).

## INTRODUCTION

Due to a lack of institutional, free stop smoking services, the role of general practitioners (GPs) in Germany is of particular importance in the treatment of tobacco addiction. However, only around one fifth of smokers in Germany who consult a GP currently receive advice to quit smoking, and an even smaller minority receives a recommendation for adequate cessation treatment (e.g., nicotine replacement therapy (NRT), bupropion and varenicline) [1]. Evidence-based strategies are therefore needed to overcome barriers preventing GPs in Germany from routinely providing brief stop-smoking advice and recommendations for evidence-based cessation treatment to their smoking patients [2-5].

As such a strategy, we developed and pre-tested two 3.5h-trainings for GPs in delivering brief stop-smoking advice [6] aiming to address the most frequently mentioned barriers towards a routine provision of stop-smoking advice: the lack of training and skills of GPs on how to provide stop-smoking advice effectively and efficiently and the lack of time during routine consultations [7-12]. One training was designed to instruct GPs how to deliver such advice according to the 5As counselling method (ask, advice, assess, assist, arrange, including the 5Rs in patients unmotivated to quit) [2], the other training to deliver the briefer ABC method (ask, brief advice, assist) [13].

In a previous pragmatic, two-arm cluster randomised controlled trial (cRCT), we showed that these trainings were highly effective at increasing the patient-reported rates of GP-delivered stop-smoking advice (primary outcome) and of delivered recommendations or prescriptions of behavioural and pharmacological smoking cessation treatment (secondary outcomes) [6 14]. A comparison of the effectiveness of the ABC vs. the 5As method regarding these primary and secondary outcomes indicated that the ABC method may lead to higher rates of GP-delivered stop-smoking advice [14].

Evidence is strong that such advice significantly increases the rates of successful quitting, in particular when combined with pharmacological treatment [15 16], and that training health professionals in advising their smoking patients can have measurable effects on continuous abstinence [17]. Hence, by increasing GP-delivered rates of advice on quitting and on effective treatment, we expected a higher number of smoking patients to intent to and to successfully quit following our GP training. However, among the few trials on the effectiveness of training particularly GPs [17-20] or GP trainees [12] so far, only the studies of Verbiest et al. [21] and Unrod et al. [20] examined the indirect effect of such a GP training on the patients’ cessation behaviour. Both studies found a positive effect of a 40min individual training [20] or respectively a one-hour 5As group training [21] on GPs counselling behaviour, but no [21] or, respectively, no clear [20] transfer of these effects on smokers’ long-term quit rates, and no effects on smokers’ attempts to quit [20 21]. Furthermore, studies conducted in the primary care context in Germany are lacking.

The present follow-up analysis of pre-specified [6], additional secondary outcomes of our above mentioned cRCT therefore examined whether our 3.5h-training for GPs has the potential to indirectly increase patients’ quit attempts and point prevalence abstinence rates at weeks 4, 12, and 26 following the consultation with the trained GP. Furthermore, we compared the ABC and 5As training regarding these outcomes.

## METHODS

### Study design and participants

The trial methods and primary outcome data are reported in detail elsewhere [6 14]. Here, we briefly summarise the main methods with a focus on follow-up data assessment. Between June 2017 and February 2020, we conducted a pragmatic, two-arm cRCT training GPs in delivering brief stop-smoking advice according to two different methods of giving advice – ABC and 5As – with a pre-post-design for the primary outcome as well as cluster randomisation for the comparison of both methods against each other. Ethics approval was obtained from the ethics committee at the Heinrich-Heine-University Düsseldorf, Germany (5999R), and all study participants provided written informed consent.

GP practices were recruited across the German federal state North Rhine-Westphalia [6 14]. A total of 52 GP practices (cluster) with 69 GPs were included in the statistical analyses of the primary and some secondary outcomes. Twenty-seven practices (32 GPs) were randomised (1:1) to receive training on the ABC method, and 25 practices (37 GPs) were randomised to receive training on the 5As [14].

Patients were eligible to participate if aged >18 years, capable to provide informed consent, if they had an appointment with a study GP in person, and were current occasional or regular tobacco smokers (e.g., cigarettes, pipe, cigars, shisha). Patients who had participated in the pre-training data collection period were ineligible to participate in the post-training period.

### Procedures (Intervention)

In 2016, we had developed and pre-tested two 3.5h-trainings for GPs in delivering brief stop-smoking advice [6] according to either the 5As [2] or the briefer ABC [13] method. The development, based on the COM-B model of behaviour [22], and pilot testing have been described in the study protocol [6]. Both trainings included peer coaching (a senior researcher together with an experienced GP), lectures on tobacco addiction, on evidence-based cessation treatment, and the respective method of giving advice (ABC or 5As), followed by intensive, standardised role plays with professional actors [6].

### Assessment of primary and main secondary outcomes at baseline (main cRCT) [14]

Data on the primary outcome (rates of GP-delivered stop-smoking advice) and on main secondary outcomes (rates of GP-delivered recommendations of behavioural counselling, NRT, varenicline or bupropion) were collected during six weeks prior to and six weeks following the GP training in smoking patients consulting these GPs, by means of questionnaire-guided, face-to-face interviews conducted by members of the study team (baseline questionnaire: osf.io/7pmr5/, translated English version: osf.io/f2p7b/).

### Assessment of secondary outcomes at follow-up: quit attempts, point prevalence abstinence

Patients who were smokers at baseline received a brief questionnaire by postal mail at 4, 12, and 26 weeks following their GP consultation (exemplary follow-up questionnaire at week 4: https://osf.io/u835j/, translated English version: https://osf.io/8uxag/). We pre-specified a time window for the postal dispatch of +/-1 week for the first follow-up, and of +/-2 weeks for the second and third follow-up. Patients who did not answer the first follow-up questionnaire received one reminder. We aimed to increase response rates by providing a small unconditional and non-financial incentive to the patients at each follow-up (e.g., a pen or some sweets).

The following secondary outcomes were collected at all three follow-up time points:

- Patient-reported rates of quit attempts since the baseline consultation with their GP.
- Patient-reported point prevalence abstinence rates since the baseline consultation with their GP, defined as the answer *“I am still not smoking”* on a question on the duration of their most recent quit attempt.

We decided not to analyse use of evidence-based stop-smoking treatments as a further secondary outcome, mainly because usage of such therapies is generally very low in Germany [23]. Assuming a decrease in response rates across all follow-up time points, the sample size was expected to be too small to draw reliable conclusions from such analyses.

In order to attenuate recall bias and to avoid overlap with multiple GP consultations or quit attempts since the baseline consultation with their GP, all questions were thoroughly phrased with reference to the respective follow-up period.

### Statistical analysis

Our study was powered to detect a clinically relevant effect of an increase in patient-reported rates of GP-delivered stop-smoking advice by at least 10% (Odds Ratio (OR) of 1.77) for the evaluation of the primary endpoint [6 14]. We did not power the study for additional secondary outcomes or loss to follow-up.

#### Analyses of the secondary outcomes collected at three follow-up time points

Both the analysis plan (v3-3: osf.io/36kpc/) and statistical code (FU_rcode_v2-4: osf.io/5tfq7/) were written and published prior to the analyses. Writing of the code was based on a blinded dataset; i.e., with the values of the outcome variables in a randomly shuffled order. All analyses were conducted using R version 4.0.3 [24].

Data were structured hierarchically in clusters (= practices), with patients within these clusters. We used mixed-effects logistic regression models to analyse the secondary outcomes: self-reported quit attempts (yes/no) and self-reported point prevalence abstinence (yes/no) at all three follow-up time points. Models were adjusted for pre-specified potential confounders measured at baseline: patients’ age, sex, level of education, and time spent with and strength of urges to smoke [25]. The group variable and its interaction with time (post-vs. pre-training) were added to the models as fixed effects to compare the ABC vs. the 5As method. Random intercepts and slopes (for the time effect) were included at the practice level. The time effect and the interaction were analysed by means of Wald-type tests (level of significance .05).

All 1,937 patients who were smokers at baseline [14] were included in an intention-to-treat analysis. Multiple imputation was used to impute missing outcome data of each follow-up as well as missing data of potential confounders by chained equations (“mice-package” [26] with 20 imputed datasets and 10 iterations for each dataset). Results across the imputation datasets were pooled using Rubin’s rules [27]. For plausibility checks, we looked at the summary statistics with minimum and maximum values, interquartile range, mean and median across all imputed datasets per outcome. We will report ranges of imputed outcomes in percent from minimum value (min) to maximum value (max) as “min-max_imputed_” exemplary for the main results presented within the results section of this manuscript.

### Adherence to the protocol

All planned analyses are reported in the study protocol [6] and in the analysis plan (https://osf.io/36kpc/). In addition to the multilevel mixed-effects logistic regression models which we ran in R using the lme4 software package, we ran sensitivity analyses using two additional software packages (GLMMadaptive and glmmTMB) to increase confidence in the results.

We ran ancillary post-hoc analyses exploring:

a. the effect of the receipt of stop-smoking advice (yes vs. no) during the baseline consultation with the GP on the secondary outcomes at each follow-up, irrespective of the time variable (pre-post training) and the group variable (ABC or 5As training),
b. the effect of the receipt of stop-smoking advice (yes vs. no) during the baseline consultation with the GP and its interaction with the time variable (post-vs. pre-training) on the secondary outcomes at each follow-up, irrespective of group variable (ABC or 5As training), and
c. the effect of the receipt of a stop-smoking advice (yes vs. no) during the baseline consultation with the GP and its interaction with the time variable (post-vs. pre-training) and with the group variable (ABC vs. 5As training) on the secondary outcomes at each follow-up.

Multilevel mixed-effects logistic regression models were also used for these analyses, including random intercepts and slopes (for receipt of advice and time; the latter only in the models for b and c) at the practice level.

## RESULTS

The flow of study participants is depicted in a previous publication of this study [14]. In brief, 1,937 smoking patients (n=1,039 were interviewed prior to and n= 898 following the GP training) were eligible to receive follow-up questionnaires. Of these patients, 770 (39.8%) answered the follow-up questionnaire at week 4, 616 (31.8%) at week 12, and 558 (28.8%) at week 26. Of these responders, around 2% (1.7% in week 4, 1.9% in week 12, and 2.3% in week 26) had missing data on the outcome “quit attempt”, and approximately 5% (3.7% in week 4, 7.1% in week 12, and 3.4% in week 26) of participants who had attempted to quit smoking, did not provide information on their smoking status at follow-up.

**Table 1** presents baseline sociodemographic and smoking characteristics of patients who participated in at least one of the three follow-up surveys (responders, n=851, 43.9%) versus those who did not participate in any follow-up (non-responders, n=1,086, 56.1%). Responders were of older age, more likely to be female, and reported the consumption of more cigarettes per day and higher urges to smoke than non-responders.

**Table 1.** Baseline characteristics of tobacco smoking patients, stratified by whether they had participated in <u>at least one</u> of the three follow-up surveys (responder) or not (non-responder).

|  | **Responder**<br>**(n = 851)** | **Non-responder**<br>**(n = 1,086)** | **Total sample**<br>**(N =1,937)** |
| --- | --- | --- | --- |
| Age, years (mean ± SD) | 50.9 ± 14.9 | 42.2 ± 15.6 | 46.1 ± 15.9 |
| Sex |  |  |  |
| Female | 59.3% (505) | 46.7% (507) | 52.3% (1,012) |
| Male | 40.7% (346) | 52.9% (575) | 47.6% (921) |
| Level of education† |  |  |  |
| High school equiv. | 22.1% (188) | 22.2% (241) | 22.2% (429) |
| Adv. techn. college equiv. | 12.9% (110) | 15.0% (163) | 14.1% (273) |
| Secondary school equiv. | 27.1% (231) | 30.4% (330) | 29.0% (561) |
| Junior high school equiv. | 36.1% (307) | 27.7% (300) | 31.3% (607) |
| No qualification | 1.8% (15) | 4.6% (50) | 3.4% (65) |
| Cigarettes/day (mean ± SD) | 14.7 ± 9.9 | 13.8 ± 9.6 | 13.8 ± 9.3 |
| Time Spent with Urges to Smoke<br>[25]¥ (mean ± SD) | 3.1 ± 1.5 | 2.8 ± 1.5 | 2.9 ± 1.5 |
| Strength of Urges to Smoke[25]¥<br>(mean ± SD) | 2.1 ± 1.0 | 2.0 ± 0.9 | 2.0 ± 0.9 |
| Motivation To Stop Smoking[33]<br>(mean ± SD)# | 3.4 ± 1.8 | 3.2 ± 1.8 | 3.3 ± 1.8 |

Data are presented as percentage (number), unless stated otherwise. Differences when calculating the total percentage can be explained by missing data on the respective variable. †German equivalents to education levels listed in table from highest to lowest: high school equivalent (“Allgemeine Hochschulreife”), advanced technical college equivalent (“Fachhochschulreife”), secondary school equivalent (“Realschulabschluss”), junior high school equivalent (“Hauptschulabschluss”), or no qualification. ¥ Both items of the Strength of Urges to Smoke Scale (SUTS) with values ranging from 0=lowest to 6=highest urges, # Items of the Motivation to Stop Smoking Scale ranging from 1=“I don’t want to stop smoking” to 7=“I really want to stop and intend to in the next month”.

### Self-reported quit attempts and point prevalence abstinence rates

Rates of attempts to quit smoking since baseline were 18.9% (min-max_imputed_: 16.2%-20.2%) at follow-up week 4, 26.7% (min-max_imputed_: 24.8%-28.3%) at week 12, and 27.3% (min-max_imputed_: 24.3%-31.1%) at week 26. **Table 2** shows that these rates did not differ significantly between the pre- and post-training group.

**Table 2.** Patient reports on secondary study outcomes (attempt to quit smoking, point prevalence abstinence) at follow-up week 4, 12, and 26, stratified by pre-post data collection period and by training method of the GP they had consulted at baseline; and associations of these outcomes with training (post vs. pre) and its interaction with the training method (ABC vs. 5As by post vs. pre); imputed data.

| Outcome (patient-reported) | Pre-training |  |  | Post-training |  |  | aOR<sub>imputed</sub><br>Post vs. Pre<br>(95%CI)<sup>†</sup> | aROR<sub>imputed</sub><br>ABC vs. 5As<sup>†</sup> by<br>Post vs. Pre<br>(95%CI) |
| --- | --- | --- | --- | --- | --- | --- | --- | --- |
|  | Pre<sub>ABC</sub> | Pre<sub>5As</sub> | Pre<sub>total</sub> | Post<sub>ABC</sub> | Post<sub>5As</sub> | Post<sub>total</sub> |  |  |
| Quit attempt at week 4 | 19.4% | 18.7% | 19.0% | 21.4% | 15.5% | 18.7% | 0.95 (0.67-1.35) | 1.17 (0.64-2.13) |
| Quit attempt at week 12 | 25.8% | 28.1% | 26.9% | 30.6% | 21.6% | 26.5% | 1.02 (0.72-1.45) | 1.12 (0.65-1.93) |
| Quit attempt at week 26 | 29.1% | 29.5% | 29.3% | 28.8% | 20.5% | 24.9% | 0.88 (0.57-1.38) | 1.07 (0.63-1.83) |
| Point prevalence abstinence at week 4 | 6.3% | 5.6% | 6.0% | 8.8% | 2.6% | 5.9% | 0.99 (0.50-1.97) | 1.64 (0.53-5.10) |
| Point prevalence abstinence at week 12 | 9.9% | 11.8% | 10.9% | 12.6% | 0.8% | 7.1% | 0.70 (0.38-1.26) | 1.71 (0.75-3.91) |
| Point prevalence abstinence at week 26 | 10.0% | 14.1% | 12.0% | 13.8% | 5.1% | 9.7% | 1.00 (0.51-1.95) | 1.51 (0.62-3.67) |

Data are presented as imputed percentages (%), adjusted Odds Ratios (aOR), and adjusted ratios of the Odds Ratios (aROR) and 95% confidence interval (95%CI) around aOR and aROR. Since multiple imputation was used, no absolute numbers are reported within this table.
<sup>†</sup>Logistic regression models with a fixed effect for time (post- vs. pre training) and random intercepts and slopes (for the time effect) for the practices; for the ABC vs. 5As comparison: the group variable (ABC vs. 5As training) and its interaction with time were added to the models as fixed effects; both models were adjusted for patients' sex, age, level of education, time spent with urges to smoke, and strength of urges to smoke (SUTS<sup>[25]</sup>).

Point prevalence abstinence rates were 6.0% (min-max_imputed_: 4.4%-8.3%) at week 4, 9.2% (min-max_imputed_: 8.7%-13.6%) at week 12, and 11.0% (min-max_imputed_: 9.7%-14.9%) at week 26, and also did not differ significantly between the pre- and post-training group (**Table 2**).

Although the relative frequencies of reported quit attempts and point prevalence abstinence rates appeared to have increased more strongly from prior to following the training in patients whose GP had been trained according to the ABC method compared to the 5As method (adjusted ratio of the OR (aROR) between 1.07 and 1.71), no statistically significant interaction could be observed for both outcomes at any of the three follow-up time points (**Table 2**).

### Ancillary analyses

Results of the post-hoc analyses a) of the effect of the receipt of a stop-smoking advice (yes vs. no) and b) its interaction with the time variable (post-vs. pre-training) are reported in **Table 3**. Data showed that patients who received an advice to quit by their GP at the baseline consultation had around two times higher odds (adjusted Odds Ratio (aOR) between 1.90 and 2.01) of reporting a quit attempt at each of the three follow-up time points than patients who did not receive such an advice. Twenty-six weeks following baseline, patients who had been advised to quit smoking at baseline also had about two times higher odds of reporting abstinence from smoking than those who had not been advised to quit (aOR=2.22, 95% Confidence Interval=1.21-4.11).

**Table 3.** Findings from ancillary analyses of patient reports on secondary study outcomes (attempt to quit smoking, point prevalence abstinence) at follow-up week 4, 12, and 26, stratified by whether the patient had received advice to quit smoking or not by the study GP they had consulted at baseline; and associations of these outcomes with the receipt of such advice (advice vs. no advice) and its interaction with the time variable (advice vs. no advice by post-vs. pre-training); imputed data.

| Outcome (patient-reported) | No advice | Advice | Pre-training |  | Post-training |  | aOR<sub>imputed</sub><br>Advice vs. No<br>advice<br>(95%CI)<sup>†</sup> | aROR<sub>imputed</sub><br>Advice<sup>†</sup> by Post<br>vs. Pre (95%CI) |
| --- | --- | --- | --- | --- | --- | --- | --- | --- |
|  |  |  | No advice | Advice | No advice | Advice |  |  |
| Quit attempt at week 4 | 16.6% | 26.6% | 18.6% | 21.7% | 13.4% | 29.2% | **1.90 (1.28-2.82)** | 1.27 (0.64-2.53) |
| Quit attempt at week 12 | 23.5% | 37.7% | 25.9% | 33.3% | 19.8% | 40.0% | **1.97 (1.32-2.92)** | 1.42 (0.71-2.83) |
| Quit attempt at week 26 | 24.1% | 38.8% | 28.3% | 35.7% | 17.6% | 40.5% | **2.01 (1.43-2.82)** | 1.34 (0.71-2.53) |
| Point prevalence abstinence<br>at week 4 | 5.7% | 6.9% | 6.1% | 5.0% | 5.0% | 7.8% | 1.49 (0.65-3.39) | 0.95 (0.27-3.35) |
| Point prevalence abstinence<br>at week 12 | 8.9% | 10.4% | 10.6% | 12.8% | 6.2% | 9.1% | 1.21 (0.63-2.33) | 1.13 (0.40-3.19) |
| Point prevalence abstinence<br>at week 26 | 9.4% | 16.7% | 10.9% | 19.0% | 7.1% | 15.4% | **2.22 (1.20-4.11)** | 1.04 (0.43-2.52) |

Data are presented as imputed percentages (%), adjusted Odds Ratios (aOR), and adjusted ratios of the Odds Ratios (aROR) and 95% confidence interval (95%CI) around aOR and aROR. Since multiple imputation was used, no absolute numbers are reported within this table; ORs printed in bold are statistically significant (p<0.05).
<sup>†</sup>Logistic regression models with a fixed effect for advice (advice vs. no advice) and random intercepts and slopes (for receipt of advice) for the practices; for the post- vs. pre-training comparison: the time variable was added to the model as a fixed and random effect; both models were adjusted for patients' sex, age, level of education, time spent with urges to smoke, and strength of urges to smoke (SUTS<sup>[23]</sup>).

Directions of the ORs suggest a larger increase of rates of quit attempts between pre- and post-training in patients who received advice to quit versus those who did not (aRORs between 1.27 and 1.42; **Table 3**). However, at none of the follow-ups were statistically significant interaction effects found – neither for attempt to quit nor for tobacco abstinence (all p >.05).

Results of the post-hoc analysis c) on the effect of the receipt of a stop-smoking advice and its three-way interaction with the time variable and the group variable (advice vs. no advice by post-vs. pre-training by ABC vs. 5As training) are not depicted in Table 3. Neither of these interaction effects was statistically significant (all p >.05). Confidence intervals were extremely wide, making it difficult to draw reliable conclusions from these results.

## DISCUSSION

In this follow-up analysis of a cRCT, we examined the effect of a 3.5h-training for GPs in delivering brief advice to quit to their smoking patients and compared two methods of giving such advice – ABC vs. 5As – on patients’ self-reported smoking cessation behaviour – attempts to quit and point prevalence abstinence rates – at weeks 4, 12, and 26 following a routine consultation with the study GP. By means of post-hoc analyses we further explored whether GPs’ advice on quitting in this study was associated with quit attempts and success of these attempts in smoking patients at follow-up.

Congruent with the existing evidence [15], post-hoc analyses demonstrated that GPs’ advice on quitting smoking (irrespective of our training) was related to a two-fold increase in the odds of patients’ attempts to quit at all follow-up time points and in self-reported, point prevalence tobacco abstinence at week 26. However, whereas our training substantially increased the rates of GP-delivered advice on quitting and recommendations of evidence-based cessation treatment (primary and secondary study outcomes published elsewhere [14]), the present follow-up analysis did not confirm a transfer of this improved GP performance on patients’ short- and long-term quitting behaviour. Although relative rates and direction of the ORs indicate a stronger increase of quit attempts and abstinence rates in patients whose GPs had been trained according to ABC compared to the 5As, neither attempts to quit nor abstinence rates differed significantly from pre- to post-training or in the ABC compared to the 5As group.

One can only speculate about the reasons for the lack of transfer of effects measured at the GPs level to the behaviour of the patients but probable explanations might exist on two levels. First, the training provides content for GPs to advice smokers to quit but probably lacks in-depth content on how to assist smokers effectively with quitting, or because GPs cannot deliver the contents effectively to their patients. While the main results of the study [14] showed a significant increase from pre- to post-training in GP-delivered recommendation rates for evidence-based cessation treatment, the relative numbers of recommendations were still very low after the training (<8%). Possibly, GPs may have reservations about the effectiveness and side-effects of stop-smoking medication. Such worries were in fact mentioned very often by the study GPs during the training and have previously been reported by other studies [e.g., 28 29]. This might have not been adequately addressed by our brief training. However, physicians’ advice to quit is most effective when combined with an offer of pharmacological or behavioural treatment [30].

Second, contextual factors might have hampered the transfer of GPs’ effective advice and support into patients’ behaviour change. For example, the low usage rates of evidence-based smoking cessation treatment may be due to the lack of reimbursement of costs for such treatments in the German healthcare setting [23]. Evidence shows that the usage and free availability of such treatments substantially increase the rates of quit attempts and success [31]. Although more patients were advised to quit following the GP training, only those who could and were willing to afford effective treatment might have attempted to quit. Since we did not collect socioeconomic data of patients, we cannot verify this hypothesis but recent population data from Germany showed that higher income is associated with higher odds of using medication to assist quit attempts [32].

In addition, limitations of the study design resulting in a small study sample at follow-up also might have influenced the results measured at the patient level.

## Limitations

First, around 70% of patients were lost to follow-up across the three follow-up time points which might have affected the statistical power to detect group differences and reduce the generalisability of our results. However, by using multiple imputation methods and intention-to-treat analysis, we applied effective measures to reduce the potential for bias compared to a complete case analysis. Second, this is a secondary analysis. In a primary analysis, smoking patients would have had to be randomised into two groups (consultation following the GP training vs. consultation prior to the GP training (“care as usual”)). Third, follow-up data were assessed by self-reports and tobacco abstinence lacked biochemical verification. Data are thus susceptible to biases such as memory effects or social desirability bias, which might lead to an under- or over-reporting of the respective outcomes. A fourth limitation is that we were not able to analyse patient-reported use of evidence-based stop-smoking treatments as a secondary outcome of our GP training due to the very low usage rates of such treatment in Germany [23] and the anticipated loss to follow-up. Such data would have provided a more detailed insight into the effectiveness of our training.

Further limitations which might have affected the results of the main cRCT also apply to the follow-up analysis including the fact that only short-term training effects on GPs behaviour were studied, that we did not video record the baseline consultation and are thus not sure whether GPs had effectively implemented the ABC or 5As method, and that the study was not designed to compare the effectiveness of both training methods directly, which could only have been done with a non-pragmatic design [1].

The major strength is the pragmatic nature of this study which was conducted in the real-world GP practice setting and during routine consultations, with outcome measures being assessed on both the GP and the patient level, the latter including long-term effects.

## Conclusions and policy implications

Receiving brief stop-smoking advice during a routine consultation with a GP has a positive effect on patients’ smoking cessation behaviour. However, our one-off 3.5-h-training session for GPs in the provision of such advice according to the ABC versus the 5As method, which substantially increased advice rates on the GP level, could not further improve patients’ attempts to quit smoking and long-term quit rates. Future training studies should more broadly take contextual factors into account which could hamper the transfer of GPs’ effective advice and support into patients’ smoking behaviour change – such as access to free evidence-based cessation treatment.

## Data Availability

The data underlying this study are third-party data (de-identified participant data) and are available to researchers on reasonable request from the corresponding author. All proposals requesting data access will need to specify how it is planned to use the data, and all proposals will need approval of the trial investigator team before data release. The study protocol [6], the statistical analysis plan (v3-3: https://osf.io/36kpc/) and statistical code (FU_rcode_v2-4: https://osf.io/5tfq7/) have been published.

## Abbreviations

5As: Ask, advice, assess, assist, arrange
ABC: Ask, brief advice, cessation support
cRCT: Cluster randomised controlled trial
CI: Confidence Interval
COM-B: Capability-Opportunity-Motivation-Behavior
GP: General practitioner
NR: Nicotine replacement therapy
OR: Odds Ratio
SD: Standard deviation
SUTS: Strength of Urges to Smoke Scale

## DECLARATIONS

## Participant consent for publication

All participants (patients and GPs) gave written informed consent prior to their inclusion in the study.

## Ethics approval

The study was approved by the medical ethics committee at the Heinrich-Heine-University Düsseldorf, Germany (5999R).

## Authorship

DK conceived the study and acquired funding for the current study together with SK and VL, and co-wrote this manuscript. SK, VL, and DK developed and conducted the GP trainings. SK coordinated all study processes, and wrote the first draft of the current manuscript. WV, statistician, prepared the statistical analysis code, and advised on all statistical analyses together with DK and SK, who conducted all analyses and interpreted the data. JH, DL, SKH, and CF were mainly involved in all study processes including recruitment, data collection, data entry and cleaning. OR, general practitioner and peer trainer, was mainly involved in the final evaluation of the training manual and the didactic methods. SW and TR gave valuable feedback at the time of designing the trial, and commented on and added to the present manuscript. All named authors contributed substantially to the manuscript and agreed on its final version. SK and DK are the guarantors. The corresponding author attests that all listed authors meet authorship criteria and that no others meeting the criteria have been omitted.

## SUPPLEMENTARY MATERIAL

### Supplementary Material 1

Exemplary original German version of the “Follow-up Questionnaire” at week 4 for patients (Follow-upSurveyWeek4_ABCII_German) can be downloaded here: osf.io/7pmr5/.

### Supplementary Material 2

Exemplary translated version of the “Follow-up Questionnaire” at week 4 for patients into English (Follow-upSurveyWeek4_ABCII_Engl) can be downloaded here: osf.io/8uxag/.

## REFERENCES

1. Kastaun S, Kotz D. Brief medical advice on smoking cessation - Results of the DEBRA study [Ärztliche Kurzberatung zur Tabakentwöhnung – Ergebnisse der DEBRA Studie]. Journal of Addiction Research and Practice [SUCHT] 2019;65:34–41. doi:10.1024/0939-5911/a000574

2. Fiore MC, Jaen CR, Baker TB, Bailey WC, Bennett G, Benowitz NL, Christiansen BA, Connell M, Curry SJ, Dorfman SF, Fraser D, Froelicher ES, Goldstein MG, Hasselblad V, Healton CG, Heishman S, Henderson PN, Heyman RB, Husten C, Koh HK, Kottke TE, Lando HA, Leitzke C, Mecklenburg RE, Mermelstein RJ, Morgan G, Mullen PD, Murray EW, Orleans CT, Piper ME, Robinson L, Stitzer ML, Theobald W, Tommasello AC, Villejo L, Wewers ME, Williams C, Treati CPG. A Clinical Practice Guideline for Treating Tobacco Use and Dependence: 2008 Update - A US Public Health Service report. Am J Prev Med 2008;35:158–76. doi:10.1016/j.amepre.2008.04.009

3. National Institute for Clinical Excellence (NICE). Smoking: acute, maternity and mental health services, Guidance PH48. 2013, https://www.nice.org.uk/guidance/ph48 (accessed 17 June 2020).

4. Association of the Scientific Medical Societies (AWMF) [Arbeitsgemeinschaft der Wissenschaftlichen Medizinischen Fachgesellschaften (AWMF)]. S3 Guideline “Smoking and Tobacco Addiction: Screening, Diagnostics, and Treatment” [S3-Leitlinie “Rauchen und Tabakabhängigkeit: Screening, Diagnostik und Behandlung”]. AWMF-Register Nr. 076-006 2021, https://www.awmf.org/leitlinien/detail/ll/076-006.html (accessed 01 February 2021).

5. Van Schayck Ocp, Williams S, Barchilon V, Baxter N, Jawad M, Katsaounou PA, Kirenga BJ, Panaitescu C, Tsiligianni IG, Zwar N, Ostrem A. Treating tobacco dependence: guidance for primary care on life-saving interventions. Position statement of the IPCRG. NPJ Prim Care Respir 2017;27:38. doi:10.1038/s41533-017-0039-5

6. Kastaun S, Leve V, Hildebrandt J, Funke C, Becker S, Lubisch D, Viechtbauer W, Reddemann O, Hempel L, McRobbie H, Raupach T, West R, Kotz D. Effectiveness of training general practitioners to improve the implementation of brief stop-smoking advice in German primary care: study protocol of a pragmatic, 2-arm cluster randomised controlled trial (the ABCII trial). BMC Fam Prac 2019;20:107. doi:10.1186/s12875-019-0986-8

7. Strobel L, Schneider NK, Krampe H, Beissbarth T, Pukrop T, Anders S, West R, Aveyard P, Raupach T. German medical students lack knowledge of how to treat smoking and problem drinking. Addiction 2012;107:1878–82. doi:10.1111/j.1360-0443.2012.03907.x

8. Twardella D, Brenner H. Lack of training as a central barrier to the promotion of smoking cessation: a survey among general practitioners in Germany. Eur J Public Health 2005;15:140–5. doi:10.1093/eurpub/cki123

9. Hoch E, Franke A, Sonntag H, Jahn B, Mühlig S, Wittchen H-U. Smoking cessation in primary medical care - chance or fiction? Results of the “Smoking and Nicotine Dependence Awareness and Screening (SNICAS)” study [Raucherentwöhnung in der primärärztlichen Versorgung – Chance oder Fiktion? Ergebnisse der “Smoking and Nicotine Dependence Awareness and Screening (SNICAS)”-Studie]. Addiction medicine in research and practice [Suchtmedizin in Forschung und Praxis] 2004;6:47-51. doi:

10. Raupach T, Merker J, Hasenfuss G, Andreas S, Pipe A. Knowledge gaps about smoking cessation in hospitalized patients and their doctors. Eur J Prev Cardiol 2011;18:334–41. doi:10.1177/1741826710389370

11. Cancer Research UK. Smoking Cessation in Primary Care: A cross-sectional survey of primary care health practitioners in the UK and the use of Very Brief Advice. 2019, https://www.cancerresearchuk.org/sites/default/files/tobacco_pc_report_to_publish_-_full12.pdf (accessed 02 June 2020).

12. Bobak A, Raupach T. Effect of a short smoking cessation training session on smoking cessation behaviour and its determinants among GP trainees in England. Nicotine Tob Res 201710.1093/ntr/ntx241

13. Ministry of Health. The New Zealand Guidelines for Helping People to Stop Smoking. 2014, http://www.health.govt.nz/publication/new-zealand-guidelines-helping-people-stop-smoking (accessed 06 June 2020).

14. Kastaun S, Leve V, Hildebrandt J, Funke C, Klosterhalfen S, Lubisch D, Reddemann O, McRobbie H, Raupach T, West R, Wilm S, Viechtbauer W, Kotz D. Training general practitioners in the ABC vs. 5As method of delivering stop-smoking advice: a pragmatic, two-arm cluster randomised controlled trial. ERJ Open Research 2020:00621-2020. doi:10.1183/23120541.00621-2020

15. Stead LF, Buitrago D, Preciado N, Sanchez G, Hartmann-Boyce J, Lancaster T. Physician advice for smoking cessation. Cochrane Database Syst Rev 201310.1002/14651858.CD000165.pub4

16. Hartmann-Boyce J, Hong B, Livingstone-Banks J, Wheat H, Fanshawe TR. Additional behavioural support as an adjunct to pharmacotherapy for smoking cessation. Cochrane Database Syst Rev 201910.1002/14651858.CD009670.pub4

17. Carson KV, Verbiest MEA, Crone MR, Brinn MP, Esterman AJ, Assendelft WJJ, Smith BJ. Training health professionals in smoking cessation. Cochrane Database Syst Rev 201210.1002/14651858.CD000214.pub2

18. Girvalaki C, Papadakis S, Vardavas C, Pipe AL, Petridou E, Tsiligianni I, Lionis C, Ti TANCP. Training General Practitioners in Evidence-Based Tobacco Treatment: An Evaluation of the Tobacco Treatment Training Network in Crete (TiTAN-Crete) Intervention. Health Educ Behav 2018:1090198118775481. doi:10.1177/1090198118775481

19. McRobbie H, Hajek P, Feder G, Eldridge S. A cluster-randomised controlled trial of a brief training session to facilitate general practitioner referral to smoking cessation treatment. Tob Control 2008;17:173–6. doi:10.1136/tc.2008.024802

20. Unrod M, Smith M, Spring B, DePue J, Redd W, Winkel G. Randomized Controlled Trial of a Computer-Based, Tailored Intervention to Increase Smoking Cessation Counseling by Primary Care Physicians. J Gen Intern Med 2007;22:478–84. doi:10.1007/s11606-006-0069-0

21. Verbiest ME, Crone MR, Scharloo M, Chavannes NH, van der Meer V, Kaptein AA, Assendelft WJ. One-hour training for general practitioners in reducing the implementation gap of smoking cessation care: a cluster-randomized controlled trial. Nicotine Tob Res 2014;16:1–10. doi:10.1093/ntr/ntt100

22. Michie S, van Stralen MM, West R. The behaviour change wheel: a new method for characterising and designing behaviour change interventions. Implement Sci 2011;6:42. doi:10.1186/1748-5908-6-42

23. Kotz D, Batra A, Kastaun S. Smoking Cessation Attempts and Common Strategies Employed. Dtsch Arztebl Int 2020;117:7–13. doi:10.3238/arztebl.2020.0007

24. R: A language and environment for statistical computing. R Foundation for Statistical Computing. 4.0.3 version. Vienna: Austria: R Core Team, 2020.

25. Fidler JA, Shahab L, West R. Strength of urges to smoke as a measure of severity of cigarette dependence: comparison with the Fagerstrom Test for Nicotine Dependence and its components. Addiction 2011;106:631–8. doi:10.1111/j.1360-0443.2010.03226.x

26. van Buuren S, Groothuis-Oudshoorn K. mice: Multivariate Imputation by Chained Equations in R. J Stat Softw 2011;45:1–67. doi:10.18637/jss.v045.i03

27. Rubin DB. Multiple Imputation for Nonresponse in Surveys. New York: John Wiley and Sons 1987.

28. Vogt F, Hall S, Marteau TM. General practitioners’ beliefs about effectiveness and intentions to prescribe smoking cessation medications: qualitative and quantitative studies. BMC Public Health 2006;6:277. doi:10.1186/1471-2458-6-277

29. van Eerd Eam, Bech Risør M, Spigt M, Godycki-Cwirko M, Andreeva E, Francis N, Wollny A, Melbye H, van Schayck O, Kotz D. Why do physicians lack engagement with smoking cessation treatment in their COPD patients? A multinational qualitative study. NPJ Prim Care Respir 2017;27:41. doi:10.1038/s41533-017-0038-6

30. Stead LF, Koilpillai P, Fanshawe TR, Lancaster T. Combined pharmacotherapy and behavioural interventions for smoking cessation. Cochrane Database Syst Rev 201610.1002/14651858.CD008286.pub3

31. van den Brand FA, Nagelhout GE, Reda AA, Winkens B, Evers S, Kotz D, van Schayck Ocp. Healthcare financing systems for increasing the use of tobacco dependence treatment. Cochrane Database Syst Rev 201710.1002/14651858.CD004305.pub5

32. Kastaun S, Brown J, Kotz D. Association between income and education with quit attempts, use of cessation aids, and short-term success in tobacco smokers: A social gradient analysis from a population-based cross-sectional household survey in Germany (DEBRA study). Addict Behav 2020;111:106553. doi:10.1016/j.addbeh.2020.106553

33. Kotz D, Brown J, West R. Predictive validity of the Motivation To Stop Scale (MTSS): a single-item measure of motivation to stop smoking. Drug Alcohol Depend 2013;128:15–9. doi:10.1016/j.drugalcdep.2012.07.012

